# Cell-Cell Communication Patterns in Alzheimer’s Disease Differ by Cortical Layer

**DOI:** 10.64898/2026.10.05.26364776

**Authors:** Qile Dai, Molly Murphy, Jingjing Lyu, Shizhen Tang, Jian Hu, Shinya Tasaki, Yanling Wang, David A. Bennett, Vilas Menon, Philip L. De Jager, Aron S. Buchman, Michael P. Epstein, Jingjing Yang

**Affiliations:** Center for Computational and Quantitative Genetics, Department of Human Genetics, Emory University School of Medicine, Atlanta, Georgia 30322, United States of America; Department of Statistics, Radiation Effects Research Foundation, Hiroshima, 732-0815, Japan; Rush Alzheimer’s Disease Center, Rush University Medical Center, Chicago, IL, 60612, USA; Center for Translational and Computational Neuroimmunology, Department of Neurology and Taub Institute for Research on Alzheimer’s Disease and the Aging Brain, Columbia University Irving Medical Center, New York, NY10032, USA

**Keywords:** Cell-cell communication, Alzheimer’s disease dementia, ADD, mild cognitive impairment, MCI, spatial transcriptomics, snRNAseq, dorsolateral prefrontal cortex, microglia, STACCato

## Abstract

**INTRODUCTION:** Existing cell-cell communication (CCC) studies of Alzheimer’s disease dementia (ADD) using cortical snRNAseq data do not account for the distinct cell-type and gene-expression profiles within cortical layers. Cortical-layer-aware CCC analysis may clarify biological mechanisms associated with ADD and mild cognitive impairment (MCI).

**METHODS:** We applied our recent analytical tool STACCato to identify CCC events associated with ADD and MCI within cortical layers, using a large-scale prefrontal cortex snRNAseq dataset (∼1.4M nuclei, n=395) with cortical layers annotated from reference spatial transcriptomics data.

**RESULTS:** We identified ADD/MCI-related CCC events with distinct patterns within cortical layers, including ADD-related interactions involving the known AD risk genes *APOE* and *APP* that were detected only in microglia in layer 5, and those with opposite effects of ADD and MCI in layers 5 and 6.

**DISCUSSION:** Our findings on cortical-layer-aware ADD/MCI-related CCC events underscore the importance of spatially-aware genomics studies of AD.

## 1 BACKGROUND

Alzheimer’s disease (AD), the most common cause of dementia, is a complex neurodegenerative disorder characterized by memory impairment and cognitive decline^1^. Genetic and genomic studies have provided valuable insights into its etiology^2–5^. Recent advances in single-nucleus RNA sequencing (snRNAseq) have enabled detailed characterization of gene-expression profiles across diverse cell types in large cohorts, creating unprecedented opportunities to investigate cell-type-specific transcriptomic mechanisms associated with AD^6–9^. In particular, snRNAseq data also support cell-cell communication (CCC) analyses that identify ligand-receptor interactions between sender and receiver cell types associated with AD^10–12^. Previous CCC studies have yielded important insights; for example, excitatory-to-inhibitory neuronal communication was increased in AD, whereas communication from inhibitory neurons and other non-neuronal cells to other cell types was decreased^12^.

Current analytical strategies for studying CCC patterns in AD have several important limitations. Because single-nucleus spatial transcriptomic data from postmortem brains remain limited, existing CCC analyses generally cannot detect spatially-aware AD-related CCC events. This limitation is important given the spatial complexity of transcriptomic profiles in the human brain, including cortical-layer dependent profiles^13,14^. Moreover, compared with superficial layers (layers 2–4), deeper cortical layers (layers 5–6) are more resistant to β-amyloid toxicity, a hallmark of AD pathology, highlighting the potential importance of layer-specific differences^15^. Accordingly, cortical-layer annotations (layers 1–6 and white matter) should be considered when studying CCC patterns in snRNAseq data from AD brains.

Beyond the omission of cortical-layer differences, current CCC studies of AD have additional limitations. In large, multi-sample, multi-condition snRNAseq datasets, existing methods often cannot account for sample-level confounders^16–20^. Most methods also do not support rigorous statistical testing of CCC events associated with conditions of interest^16–20^, such as the presence or absence of ADD. Finally, few studies have examined CCC events in mild cognitive impairment (MCI)^10–12^, an intermediate stage between normal cognitive aging and ADD under National Institute on Aging–Alzheimer’s Association clinical criteria, in which cognitive impair is more severe than expected but some cognitive function is still retained^21,22^. Although some individuals with MCI may never progress to dementia in their life time, or revert to normal cognition at one time point in time, individuals with MCI do face an elevated risk of progressing to dementia^23^. MCI is an important focus for investigating early ADD mechanisms and potential interventions^24,25^.

To address these gaps, we used a large-scale snRNAseq dataset from dorsolateral prefrontal cortex tissue (∼1.4M nuclei) of 395 postmortem donors in the Religious Orders Study and Rush Memory and Aging Project (ROS/MAP) cohorts^7,26^, together with detailed ADD/MCI phenotypes, demographic variables, and clinical variables^27,28^. In our previous study^9^, we used the deep-learning tool CeLEry^29^ and reference prefrontal cortex spatial transcriptomic data from the Lieber Institute for Brain Development (LIBD) project^30^ to predict cortical-layer annotations (layers 1–6 and white matter) for these nuclei. In this study, we used our recently developed analytical tool STACCato (that implements a supervised tensor regression model)^31^ to test for ADD- and MCI-related CCC events within each predicted cortical layer, while adjusting for potential sample-level confounders. We found distinct patterns of putative ADD/MCI related CCC events across layers, especially in layers 5 and 6. For example, in layer 5, the detected putative ADD-related CCC events, involving microglia and other cell types and having positive effects for ADD status, were more than those in the other layers. In layers 5 and 6, a set of putative ADD and MCI related CCC events were found with opposite effect size directions for ADD and MCI statuses.

## 2 METHODS

### 2.1 Overview

We sought to identify CCC events associated with ADD or MCI within each cortical layer (layer-dependent analyses) and compare them with findings obtained without spatial stratification (layer-independent analysis). Layer-dependent analyses included only nuclei annotated to a target cortical layer (1-6) or white matter (WM), whereas the layer-independent analysis included all nuclei across the six cortical layers and WM. We applied the same preprocessing and CCC analysis procedures to each layer-specific subset and to the complete ROS/MAP snRNAseq dataset. We considered six major brain cell types: astrocytes, excitatory neurons, inhibitory neurons, microglia, oligodendrocytes, and oligodendrocyte precursor cells (OPCs). For each analysis, we used our recently developed analytical tool STACCato^31^ to identify ADD- and MCI-related CCC events while adjusting for potential sample-level confounders, including age, sex, education, *APOE E4* and *TOMM40* variants, and recent medication use.

### 2.2 ROS/MAP snRNAseq data

The ROS/MAP dataset comprises harmonized data from two large, prospective, longitudinal, community-based clinicopathological cohort studies, ROS and MAP^32–34^, which use common protocols employed by the same staff^28^. Participants were enrolled without known dementia and underwent annual assessments of diverse clinical covariates and cognitive status. Participants consented to brain donation after death, and both studies followed protocols approved by an Institutional Review Board of Rush University Medical Center. Each participant provided informed and repository consent and signed an Anatomical Gift Act.

The ROS/MAP snRNAseq dataset comprises transcriptomic profiles from ∼1.4M cells obtained from 424 postmortem DLPFC tissue samples^7,26^. We analyzed 395 samples with complete demographic and clinical data, including summary clinical cognitive diagnosis, age at death, sex, education, *TOMM40* haplotypes, *APOE E4* genotypes, and recent use of ADD, lipid-lowering, and urinary-condition medications within the preceding two weeks. Based on the summary clinical cognitive diagnosis closest to death, participants were classified as having no cognitive impairment (NCI; n=138), MCI (n=107), or ADD (n=150)

### 2.3 Cortical layer annotation and quality-control preprocessing of ROS/MAP snRNAseq data

Our previous study used CeLEry^29^ and LIBD prefrontal cortex spatial transcriptomic data^30^ as a reference to predict cortical-layer annotations (layers 1–6 and WM) for all nuclei in the ROS/MAP snRNAseq dataset^9^. CeLEry trains a deep-learning model to learn relationships between gene-expression profiles and spatial locations in a reference spatial transcriptomic dataset and then predicts cell locations from snRNAseq expression profiles^29^. The proportions of six major cell types (astrocytes, excitatory neurons, inhibitory neurons, microglia, oligodendrocytes, and OPCs) in each inferred cortical layer were validated against a single-cell-resolution spatial transcriptomic dataset of prefrontal cortex^9^.

We then performed quality control (QC) and preprocessing for each layer-dependent ROS/MAP snRNAseq subset and the overall layer-independent dataset, as detailed in the Supplementary Methods. **Table S1** summarizes the numbers of ADD, MCI, and NCI samples retained after QC for each subset and full dataset. The layer-independent analysis included all 395 samples with complete summary clinical cognitive diagnoses and covariates. Analyses of layers 2, 3, and 4 also included all 395 samples. Analyses of layers 1, 5, 6, and WM excluded 7, 5, 7, and 12 samples, respectively, because those samples contained only one cell type.

### 2.4 STACCato analysis for detecting putative ADD- and MCI- related CCC events

After QC, we quantified the snRNAseq data as pseudo bulk read counts (average read counts across cells) per gene per sample, normalized by scaling library size factors to 10^4^, and then subjected these counts to a log-transformation. Using these log-transformed read counts, we applied LIANA+^35^ to construct CCC scores for the consensus list of ligand-receptor pairs used by eight existing CCC analysis tools (e.g., CellChat^16^, NATMI^18^, SingleCellSigmalR^19^, and CellPhoneDB^36^), which are curated from OmniPath^37^. Using the mean scoring function (***Equation S1***) proposed by CellPhoneDB^36^, we calculated CCC scores for each sample in the snRNAseq data across the consensus set of ligand-receptor pairs and all pair-wise combinations of sender and receiver cell types. The ligand-receptor directions are determined by known annotations from the OmniPath^37^. The CCC scores are determined by the gene expression levels in the genes of the corresponding ligand-receptor pair in the each pair of sender and receiver cell types.

We aggregated the CCC scores into a four-dimensional tensor representing samples, ligand-receptor pairs, sender cell types, and receiver cell types, respectively in its four dimensions. We then applied the STACCato tool^31^ to detect CCC events associated with ADD or MCI while adjusting for age, sex, education, *APOE E4* and *TOMM40* variants, and recent medication use, by using a tensor regression model. STACCato estimates the effect sizes of cognitive statuses (ADD or MCI) on each CCC event, while adjusting for covariates in the same tensor regression model. Further, STACCato provides resampling-based p-values for the estimated effect sizes (see **Supplementary Methods**). We prioritized events with nominal resampling-based p-values < 0.05 and effect-size magnitudes in the top 5% as putative disease-related CCC events.

### 2.5 Compare identified putative ADD/MCI-related CCC events across cortical layers and WM

We used STACCato for layer-dependent analyses that were restricted to cells in each cortical layer (1-6, WM), and a layer-independent analysis using the data of all cells. We then used the Jaccard index^38,39^ to quantify similarity between sets of prioritized putative ADD/MCI-related CCC events across cortical layers, and between each layer-dependent and layer-independent analyses. The Jaccard index is the size of the intersection of two sets of CCC events (A∩B) divided by the size of their union 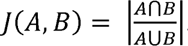, Here, A and B denote the sets identified in two distinct analyses. Higher Jaccard index values indicate greater overlap between the sets.

## 3 RESULTS

### 3.1 Cell distributions across inferred cortical layers and WM

We examined cell counts and cell-type proportions across the six inferred cortical layers and WM in the ROS/MAP snRNAseq data (**Figure 1A**). Layer 3 had the highest median cell count (884 cells per sample), followed by layer 4 (614), layer 5 (490), layer 2 (487), layer 6 (314), WM (306), and layer 1 (276). We next examined cell-type proportions across the six layers and WM (**Figure 1B**). Astrocytes were predominant in layer 1 but decreased progressively in deeper layers, whereas the proportion of oligodendrocytes increased from layer 1 to WM. Excitatory neurons were the most abundant cell type in layers 2–5 and the second most abundant in the remaining layers; their proportions increased from layer 1 to layer 5 and then declined toward WM. Inhibitory-neuron proportions increased from layer 1 to layer 3 and then declined toward WM. Microglia and oligodendrocyte precursor cells had lower overall proportions than the other cell types; microglia were more abundant in layers 1–4, whereas oligodendrocyte precursor cells were relatively enriched in layers 3–6.

**Figure 1.**
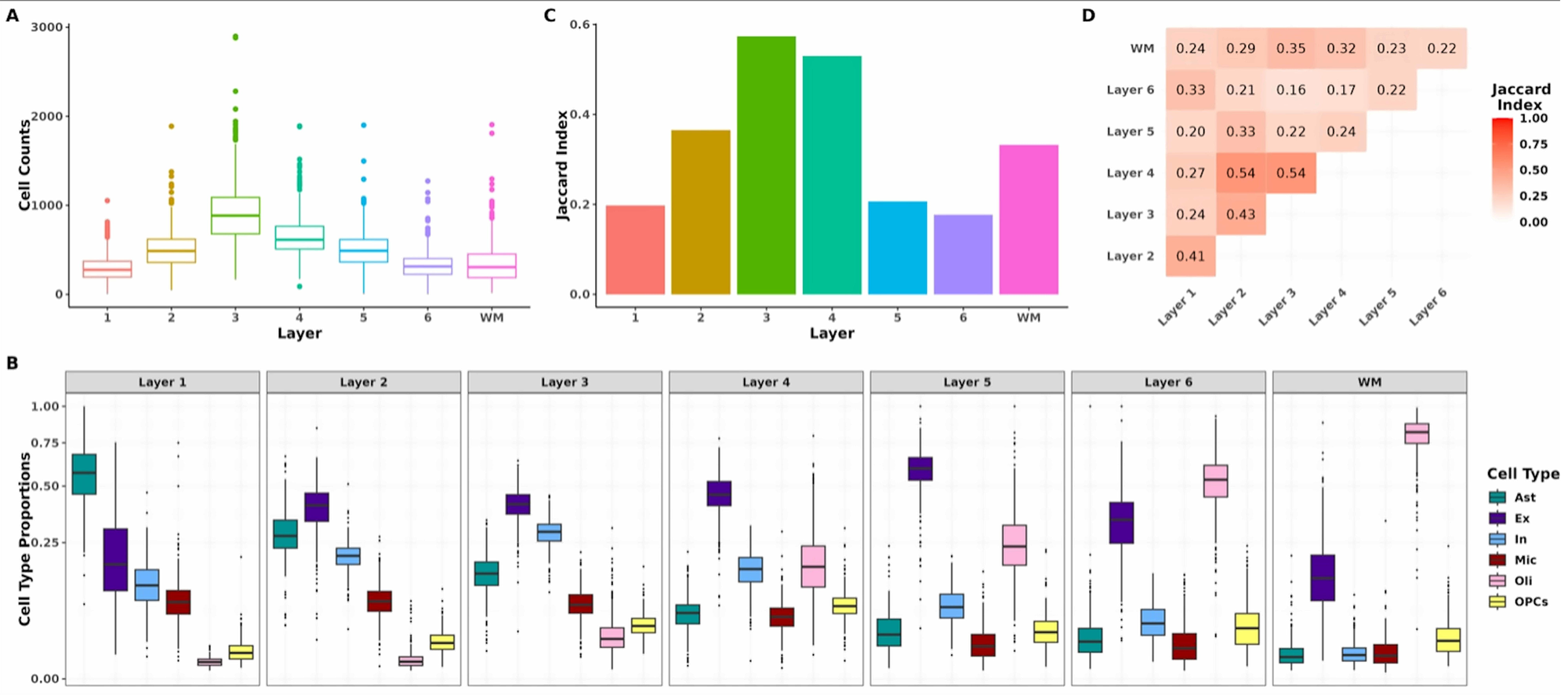
(A) Box plot of cell counts per sample across cortical layers. (B) Box plot of cell type proportions across cortical layers. (C) Bar plot quantifying the Jaccard Index for the overlap of detected putative ADD-related CCC events between layer-independent and each layer-dependent results. (D) Heatmap of the Jaccard Index to show the overlap of ADD-related CCC events between each pair of layer-dependent results. In the box plot, the box represents the interquartile range, the central line indicates the median, and the whiskers extend to the data range excluding outliers, which are shown as individual points. Abbreviations: white matter (WM), astrocytes (Ast), excitatory neurons (Ex), inhibitory neurons (In), microglia (Mic), oligodendrocytes (Oli), and oligodendrocyte precursor cells (OPCs).

### 3.2 Similarity of putative ADD-related CCC events across layers

**Table S1** reports the numbers of prioritized putative ADD-related CCC events and total analyzed CCC events for each layer-dependent analysis as well as the layer-independent results. Each analysis yielded approximately 2,000 putative (top 5%) ADD-related events among approximately 40,000 total CCC events (**Figure S1**). We used the Jaccard index^40^ to compare layer-dependent putative ADD/MCI related events with those from the layer-independent analysis (see Methods). Layers 3 and 4 had the highest Jaccard index values (**Figure 1C**), indicating the greatest similarity to the layer-independent results, whereas layer 1, layer 5, and layer 6 had lower values. Because layer 3 and layer 4 contained the largest numbers of cells, particularly neurons (**Figures 1A** and **1B**), neuronal signals from these layers may have contributed disproportionately to the layer-independent results.

To further evaluate the similarity of putative ADD-related CCC events across layers, we calculated pairwise Jaccard Index values between layers (**Figure 1D**). Some spatially adjacent layers, including layers 1 and 2, layers 2 and 3, and layers 3 and 4, exhibited higher overlaps, with layers 2 and 4 and layers 3 and 4 showing the greatest similarity. In contrast, deeper adjacent layers, such as layers 5 and 6, exhibited much lower overlap, indicating that spatial proximity does not necessarily correspond to similarity in CCC patterns. Overall, layers 5, 6, and WM were identified as having the most distinct CCC patterns based on Jaccard Index values in comparisons with all other layers.

### 3.3 Top putative ADD-related CCC events can vary by cortical layer

In the layer-independent analysis, we examined the 50 putative CCC events with the largest positive ADD-associated effects (top left panel of **Figure 2A**) and the top 50 with the largest negative effects (top left panel of **Figure 2B**). Effect directions in layer-dependent analyses were generally consistent with those in the layer-independent analysis, although magnitudes often differed. Notable exceptions included interactions between *NRXN1* in astrocytes, excitatory neurons, or inhibitory neurons and *NLGN1* in microglia, which had large negative effects in the layer-independent analysis but positive effects in layers 1 and 5 (**Figure 2B**). These findings suggest that even among the top CCC events identified by the layer-independent analysis, the functional roles of CCC events within individual layers can be different. This underscores the importance of layer-dependent analyses for uncovering the accurate functional roles of CCC events.

**Figure 2.**
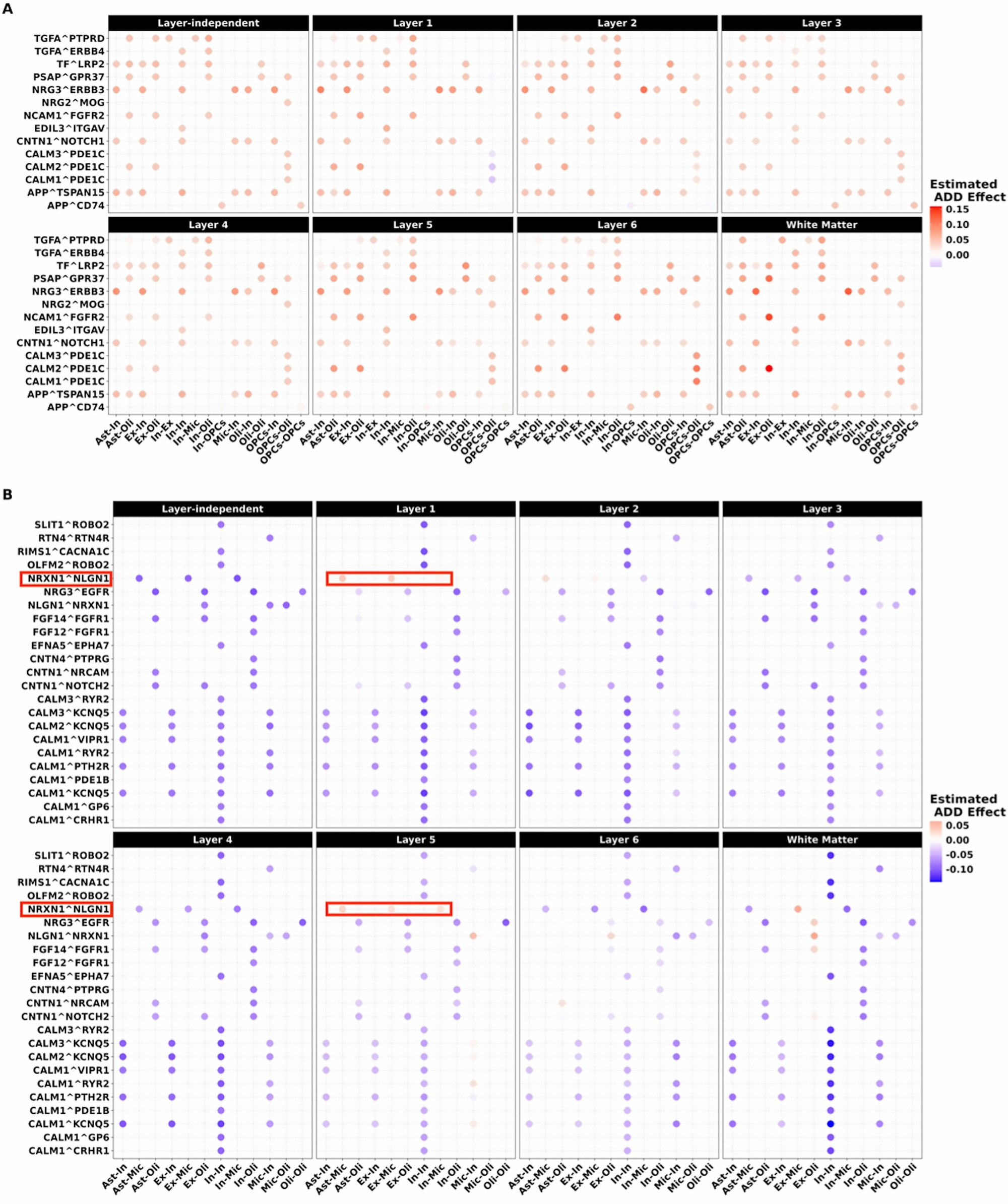
Estimated ADD disease effect on the top 50 putative CCC events with (A) largest positive magnitude and (B) largest negative magnitude in layer-independent results. Estimated ADD disease effects for these CCC events are compared between layer-independent and layer-dependent results. Positive disease effects, shown in red, indicate positive associations between CCC events and ADD, while negative disease effects, shown in blue, indicate negative associations. Within each panel, each grid represents the estimated ADD disease effect for a specific CCC event, with the ligand-receptor pair labeled on the y-axis and the sender-receiver cell type pair indicated on the x-axis. Abbreviations: astrocytes (Ast), excitatory neurons (Ex), inhibitory neurons (In), microglia (Mic), oligodendrocytes (Oli), and oligodendrocyte precursor cells (OPCs).

To further characterize layer-specific ADD-related CCC patterns, we examined the cell types involved in the putative ADD-related events by layer-independent and layer-dependent analyses (**Figure 3**). The layer-independent analysis identified substantially more negative (1,750) than positive (123) ADD-related effects (**Figure 3A**). Cell-type involvement was generally balanced across negative and positive interactions, except that microglia were minimally involved in positive associations. Layer 3 (**Figure 3D**) and layer 4 (**Figure 3E**) most closely resembled the layer-independent pattern, although layer 3 had more positive interactions involving microglia and layer 4 had more positive interactions involving oligodendrocytes. Even though layers 1 & 2 (Figure 3B & 3C) also have a greater number of negative interactions than positive ones, a larger proportion of negative interactions involving ligands in OPCs in layer 1 and a larger proportion of positive interactions involving ligands in microglia cells in layer 2 were observed.

**Figure 3.**
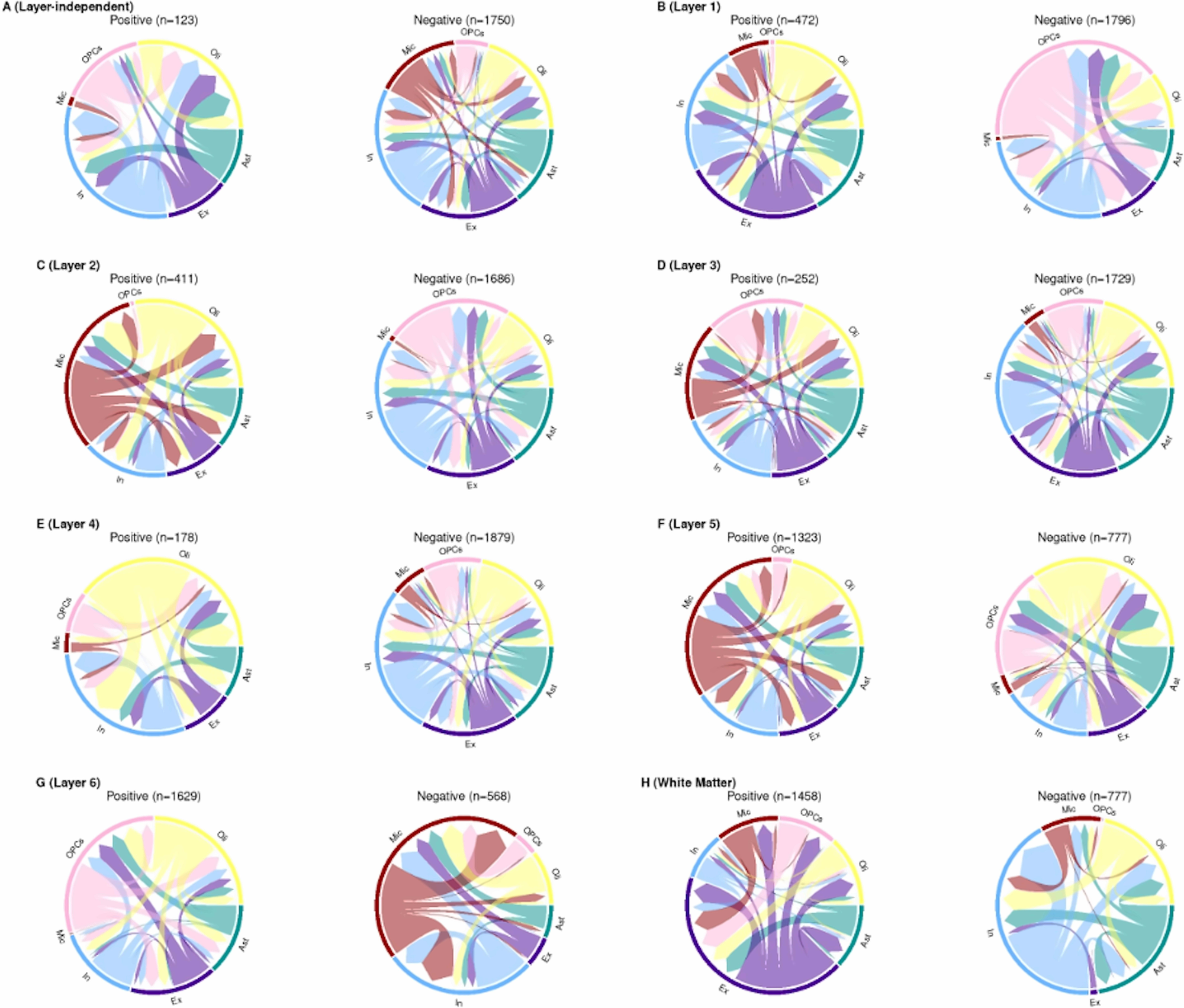
Circos plots of ADD-related communication network between cell types identified from. (A) layer-independent analysis, (B) Layer 1, (C) Layer 2, (D) Layer 3, (E) Layer 4, (F) Layer 5, (G) Layer 6, and (H) white matter. The width of the links represents the total number of detected putative ADD-related CCC events between each pair of sender-receiver cell types. The number of identified positive ADD-related CCC events are shown on the left side of each panel, while the negative events are on the right. Each color represents a different cell type. Abbreviations: astrocytes (Ast), excitatory neurons (Ex), inhibitory neurons (In), microglia (Mic), oligodendrocytes (Oli), and oligodendrocyte precursor cells (OPCs).

In contrast to layers 3 and 4, layer 5 (**Figure 3F**), layer 6 (**Figure 3G**), and WM (**Figure 3H**) exhibited communication patterns more distinct from the layer-independent results, particularly involving a substantially larger number of positive interactions than negative ones. Layer 5 (**Figure 3F**) had 1,323 positive ADD-related CCC events and 777 negative events, with microglia prominently involved as both sender and receiver cell type. Layer 6 (**Figure 3G**) had 1,629 positive and 568 negative ADD-related CCC events, with oligodendrocyte precursor cells and oligodendrocytes prominently involved as both sender and receiver cell type. WM (**Figure 3H**) showed 1,458 positive and 777 negative ADD-related CCC events, with excitatory neurons playing the most dominant role in positive events. These results highlight the importance of incorporating spatial information within genomic studies of AD.

### 3.4 Unique patterns of putative ADD-related CCC events in layer 5 involving microglia

Motivated by the distinct role of microglia in layer 5 (**Figure 3F**), we analyzed the estimated ADD effects on detected putative ADD-related CCC events within this layer, focusing specifically on events where microglia are involved (acting as either the sender or receiver cell type). The detected putative ADD effects on these CCC events in layer 5 were compared with those detected by layer-independent analyses and in the neighboring layers, layer 4 and layer 6, with **Figure 4** highlighting CCC events involving ligands or receptors encoded by known risk genes in the NHGRI-EBI genome-wide association study (GWAS) catalog^41^ with the associated trait as ‘Alzheimer disease’, while **Figures S2** and **S3** present results for other CCC events.

**Figure 4.**
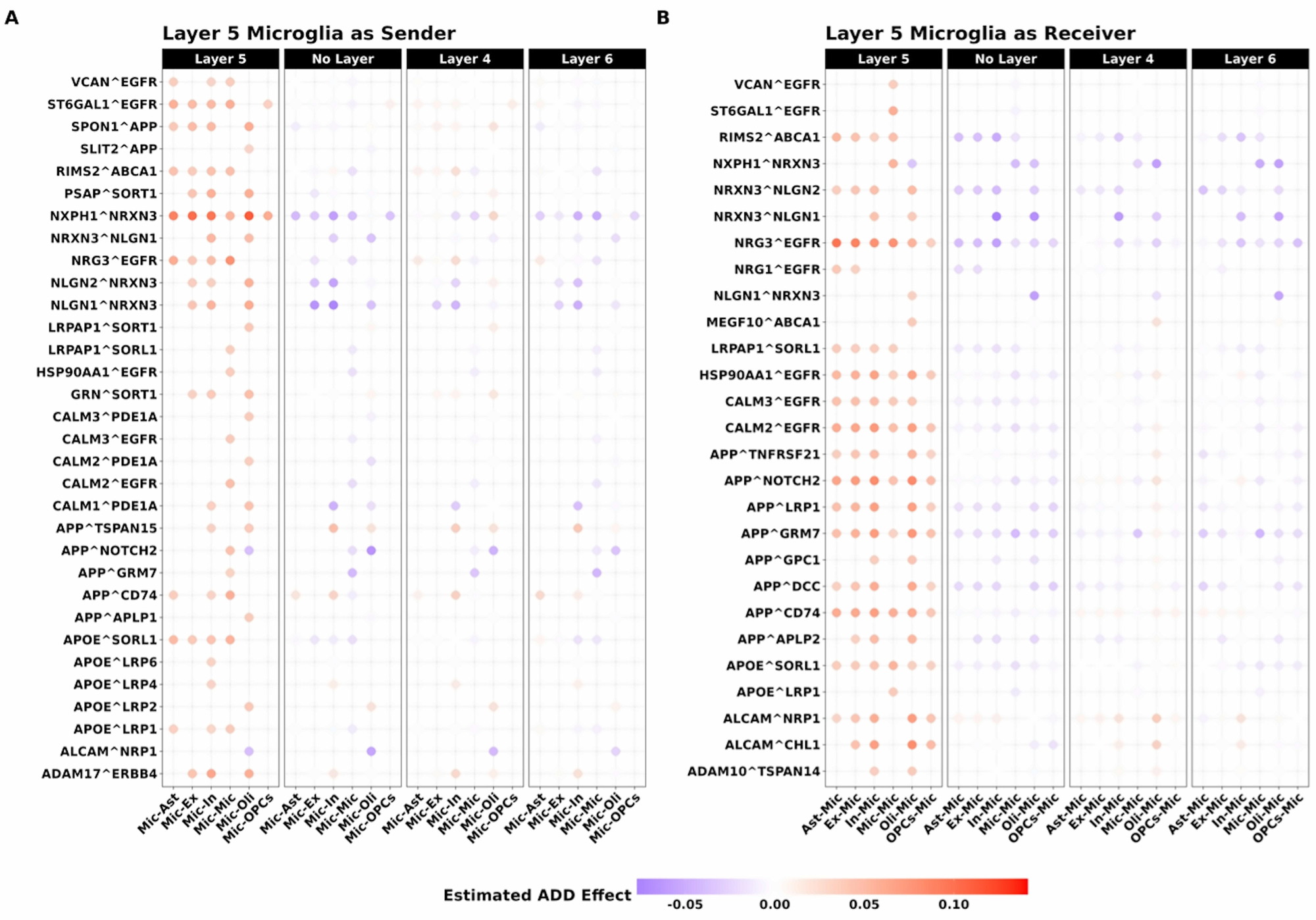
Estimated ADD disease effects on putative CCC events in Layer 5, where (A) microglia act as the sender cell type, or (B) microglia act as the receiver cell type, and the ligand-receptor pairs involve genes associated with ADD in the GWAS catalog. The estimated ADD disease effects for these CCC events are compared between Layer 5 results and layer-independent results, as well as Layer 4 and Layer 6 results. Positive disease effects, shown in red, indicate positive associations between CCC events and ADD, while negative disease effects, shown in blue, indicate negative associations. Within each panel, each grid represents the estimated ADD disease effect for a specific CCC event, with the ligand-receptor pair labeled on the y-axis and the sender-receiver cell type pair indicated on the x-axis. Abbreviations: astrocytes (Ast), excitatory neurons (Ex), inhibitory neurons (In), microglia (Mic), oligodendrocytes (Oli), and oligodendrocyte precursor cells (OPCs), genome-wide association study (GWAS).

Most detected putative ADD-related events involving microglia as sender or receiver in layer 5 were positively associated with ADD (column 1 in **Figures 4A** and **4B**; **Figures S2** and **S3**). For these events, estimated effects in the layer-independent analysis and in layers 4 and 6 were often negative or near zero (columns 2–3 in **Figures 4A** and **4B**), indicating that the observed associations were specific to layer 5. The smaller set of negatively associated events generally showed more consistent effects across the layer-independent analysis and layers 4–6.

Many positive putative ADD-related CCC events involving microglia in layer 5 contained genes that are well-studied in ADD and other neurodegenerative disorders. One such gene was *APOE*, which is widely recognized as the most significant genetic factor influencing the risk of sporadic ADD^42^. We observed several positive ADD-related CCC events involving ligand *APOE* and receptor *SORL1*; either when microglia was the sender cell type (**Figure 4A**) or the receiver cell type (**Figure 4B**). These ADD-related effects were unique to layer 5 as analogous effects in the layer-independent analysis and layers 4 and 6 were often negative or close to zero (columns 2-3 in **Figure 4A** and **4B**). In addition to *SORL1*, we also observed positive ADD-related effects of ligand *APOE* in microglia sender cells in layer 5 with low-density lipoprotein (LDL) receptor-related proteins (LRPs), including *LRP1*, *LRP2*, *LRP4*, *LRP6*, in multiple receiver cell types (column 1 in **Figure 4A**). These effects were not observed in layer-independent analysis nor analyses within layers 4 or 6.

Beyond *APOE*, several well-studied ADD-related genes showed distinct CCC patterns involving microglia in layer 5. *APP* is central to ADD pathology because its cleavage produces amyloid-beta peptides, a hallmark of *ADD*^43^. Interactions involving *APP* and *TNFRSF21, NOTCH2, LRP1, GRM7, GPC1, DCC, CD74*, or *APLP2* across microglia and other cell types were positively associated with ADD only in layer 5 (column 1 in **Figures 4A** and **4B**). Although no causal role in ADD progression can be determined from CCC analyses conducted by STACCato, these layer-specific associations suggest that microglia-related signaling in layer 5 warrants further investigation.

### 3.5 Putative MCI-related CCC events with consistent effects across cortical layers and WM

Comparing cortical layer-dependent putative MCI-related CCC events to layer-independent events (**Figure S4; Figure S5A**), we observed that the results of layer 3 had the greatest similarity (largest Jaccard Index) to layer-independent analysis, followed by layers 2, 4, and 5. We also observed that layers 1 through 6 were notably similar to their neighboring layers and the WM layer was most distinct from others (**Figure S5B**) with the smallest observed Jaccard Indexes.

In the layer-independent analysis, 1,864 putative MCI-related negative interactions were identified, greatly exceeding the 9 positive interactions (**Figure S6A**). Among these negative interactions, cell type involvement was relatively balanced, with inhibitory neurons, OPCs, and microglia participating slightly more frequently than other cell types. Layers 2 (**Figure S6C**), 3 (**Figure S6D**), 4 (**Figure S6E**), 5 (**Figure S6F**), and 6 (**Figure S6G**) all showed substantial similarity to the layer-independent results. Layer 1 (**Figure S6B**) also showed more negative than positive interactions, but these negative interactions were uniquely dominated by CCC events involving OPCs. In contrast, WM (**Figure S6H**) exhibited 563 putative positive MCI-related interactions, a number substantially higher than those observed in other cortical layers and the layer-independent results. Overall, compared to ADD-related CCC events, MCI-related CCC events showed greater consistency across layers and with the layer-independent results.

### 3.6 Comparison of ADD and MCI effects on CCC events

We compared estimated ADD and MCI effects across cortical layers and in the layer-independent analysis. Most CCC events had concordant directions and similar effect sizes for MCI and ADD, but some differed in direction or magnitude, particularly in layers 5 and 6 (**Figure 5**). **Figure S7** highlights ADD/MCI related CCC events with opposite ADD and MCI effect directions in these layers; such events were more numerous in layer 6 than in layer 5. Notably, compared to layer 5, layer 6 exhibits a larger number of such events with opposite directions in ADD and MCI (243 vs. 53). Among these putative CCC events with opposite directions of ADD and MCI effects, multiple ones involve *APP*, CALM family of genes (*CALM1*, *CALM2*, *CALM3*), CNTN family of genes (*CTNT1*, *CNTN2*, *CNTN4*), NLGN family of genes (*NLGN1*, *NLGN2*), and NRXN family of genes (*NRXN1*, *NRXN2*, *NRXN3*). Interestingly, all these CCC events demonstrate negative associations with MCI risk but positive associations with ADD risk, suggesting that their roles in cellular communication might shift in different disease stages. Because these analyses are cross-sectional, the contrasting associations should not be interpreted as evidence that the interactions transition from protective in MCI to pathological states in ADD.

**Figure 5.**
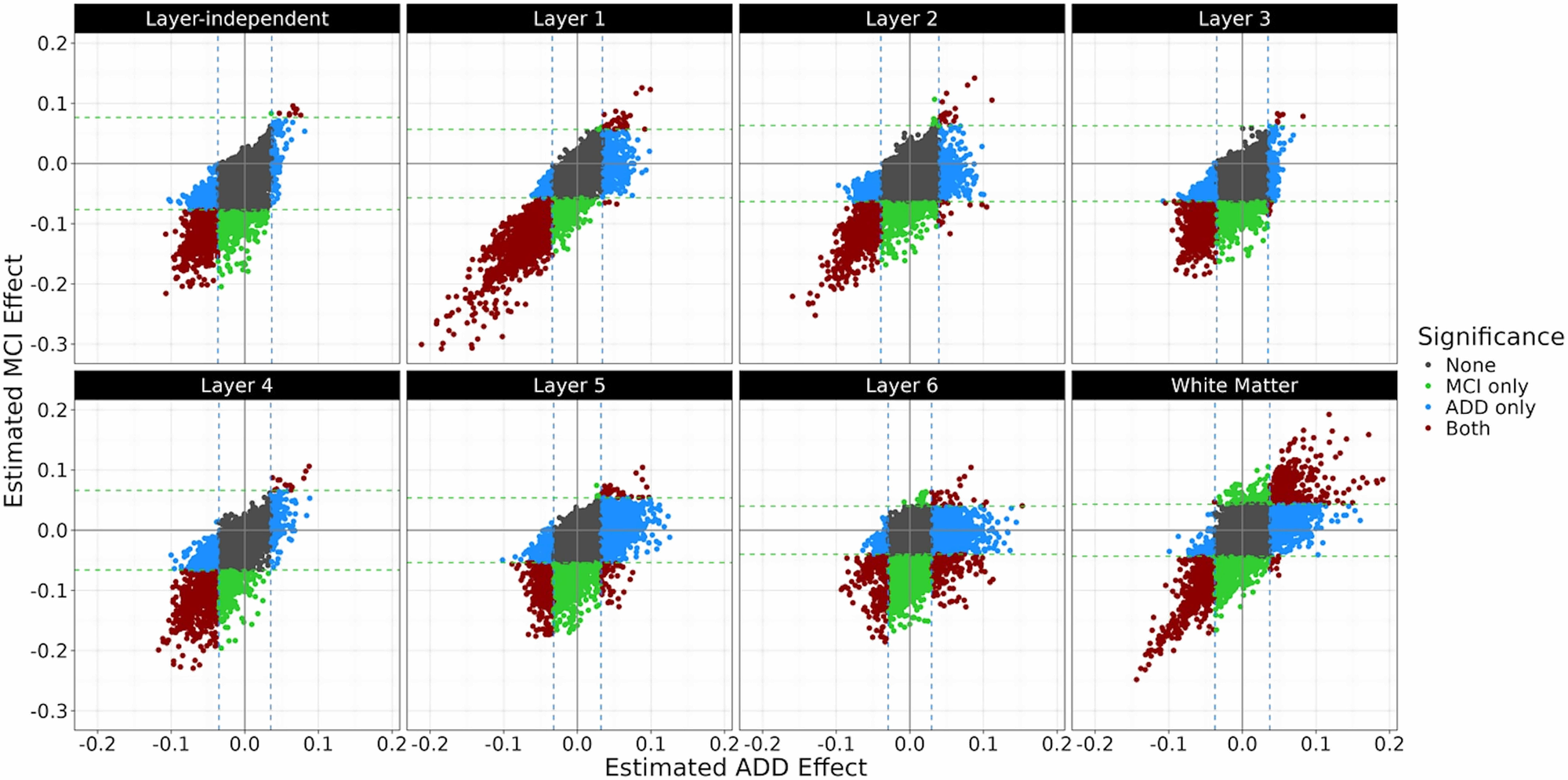
Scatter plots comparing the estimated ADD disease effects (x-axis) and MCI effects (y-axis) across different cortical layers and layer-independent results. Each point represents a CCC event, with color indicating significance: black (None) for non-significant events, red (Both) for events associated with both ADD and MCI, blue for events associated with ADD only (ADD only), and green (MCI only) for events associated with MCI only. Vertical blue dashed lines and horizontal green dashed lines represent the significance thresholds for ADD and MCI effects, respectively.

To evaluate the robustness of these ADD and MCI related CCC events with opposite effect size directions, we respectively applied STACCato to the sub-cohort of NCI and MCI participants, and the sub-cohort of NCI and ADD participants, with one binary disease status variable and the same set of confounding variables as considered in the full cohort analyses. Among these CCC events with opposite effect sizes for ADD and MCI statuses in the full cohort analysis, almost all were still estimated with the same effect size directions and similar effect size magnitudes in the sub-cohort analyses (e.g., all 53 in layer 5 and 235/243 in layer 6; **Figure S8**).

## 4 DISCUSSION

Although snRNAseq studies have successfully characterized cell type-specific biological mechanisms associated with AD^5,6,26^ through CCC analysis, spatially informed analyses remain limited because large-scale human brain spatial transcriptomics data are difficult to obtain. We addressed this gap by performing spatially informed CCC analyses using large-scale ROS/MAP snRNAseq data from prefrontal cortex tissue with cell cortical layers (layers 1-6 and WM) that were inferred from reference spatial transcriptomics data from the same tissue^9^. Reference spatial transcriptomics data therefore provided inferred layer context in the ROS/MAP snRNAseq data, rather than direct spatial measurements in the ROS/MAP samples. We aim to identify CCC events associated with ADD and MCI within each cortical layer, and compare these with the results obtained by not accounting for spatial cortical layer information.

We observed substantial variability in cell-type proportions and cell counts across inferred layers. Previous CCC analyses often pooled samples within each condition into a single pseudo-sample and compared pseudo-samples across conditions^18,44^. Although convenient, this approach assumes within-condition uniformity and obscures sample-specific variability. We instead used our recently developed analytical tool STACCato^31^, implementing a tensor regression model^45^, to both estimate effect sizes of ADD and MCI status on snRNAseq-based CCC scores and further test whether such effect sizes significantly differ from the null accounting for potential sample-level confounders (including age, gender, education level, and medication use). We note these estimates represent disease associations and not causal effects.

We compared layer-dependent results with layer-independent analyses that pooled all cells without considering inferred cortical-layer information. Layer-independent results were dominated by layers 3 and 4, which contained most cells, primarily neurons, whereas distinct patterns within individual layers and less abundant cell types were often masked. Notable layer-specific ADD/MCI related CCC events were found for those involving microglia in layer 5, OPCs and oligodendrocytes in layer 6, and excitatory neurons in the WM, which were underrepresented in layer-independent analyses. Many detected putative ADD/MCI related CCC events also had opposite estimated effect directions in specific layers and in the layer-independent analysis, indicating that their disease associations may differ across inferred cortical layers. Thus, analyses that omit inferred layer context may provide an incomplete, false positive, or misleading account of putative intercellular communication in the prefrontal cortex.

We identified numerous putative ADD/MCI related microglial interactions involving established ADD-related genes, including *APOE* and *APP*, that were uniquely associated with ADD risk in inferred layer 5. Ligand-receptor annotations from existing databases provided sender-receiver orientations and all pair-wise cell types were considered in both sender-receiver orientations in the analysis. These CCC analysis results do not establish microglia-driven signaling or a role in ADD progression. For example, *APOE-SORL1* interactions were positively associated with ADD risk. *APOE* is upregulated in neurodegenerative microglia^46,47^ and may impair the microglial response in Alzheimer’s disease^48^.

Single-nucleotide polymorphisms (SNPs) at the *SORL1* locus have also been associated with ADD across multiple ancestries^49–51^, and *APOE-SORL1* interactions have been identified as a risk factor for ADD^52,53^. Our findings refine the spatial context of these reports by localizing the association between *APOE-SORL1* CCC scores and ADD to microglia assigned to cortical layer 5.

*APOE* interactions with *LRP*s, including *LRP1, LRP2, LRP4*, and *LRP6*, were also positively associated with ADD risk in microglia annotated to cortical layer 5. Studies of *APOE* and *LRP* expression in ADD brains have reported variable and sometimes contradictory findings^54^. One study observed reduced LRP levels in the mid-frontal regions of ADD brains^55^, whereas another reported markedly increased APOE and LRP levels in the ADD frontal cortex^56^. Our results indicate that associations between *APOE-LRP* CCC scores and ADD risk may vary across inferred cortical layers, offering one possible context for these discrepancies without establishing causality.

Compared with ADD-related CCC events, MCI-related patterns were more consistent across cortical layers, whereas ADD associations were more layer specific. Layer 5 showed the most distinct pattern of putative ADD-related CCC events, indicating a particularly differentiated association profile in this layer. Most CCC events associated with both ADD and MCI had concordant effect directions; however, a subset in layers 5 and 6 was negatively associated with MCI risk and positively associated with ADD risk. These cross-sectional differences identify stage-associated contrasts in deeper cortical layers but do not demonstrate temporal change, progression, or a transition from protective effects in MCI to pathological effects in ADD.

This study has several limitations. First, cortical-layer annotations for cells in the snRNAseq dataset were inferred from reference spatial transcriptomics data of the same prefrontal cortex tissue, and uncertainty in those assignments was not propagated into the CCC analyses by STACCato. Validation in single-cell-resolution spatial transcriptomics data from ROSMAP samples and in independent snRNAseq datasets would strengthen these findings. Second, the CCC scores are calculated from gene expressions quantified by snRNAseq data and do not measure protein secretion, receptor engagement, or downstream activity. Results also depend on the completeness of curated ligand-receptor databases and the accuracy of ligand-receptor and cell-type annotations. CCC events between pair-wise cell types (with all sender-receiver orientations examined) support only putative ADD/MCI associations, which cannot establish direct physical proximity, observed communication, or signaling within or between layers. Third, inter-layer communication was not modeled. Finally, the cross-sectional postmortem design precludes causal or temporal inference, including conclusions about disease progression or protective-to-pathological transitions.

This study highlights the value of spatially informed analyses for identifying CCC events associated with ADD/MCI in postmortem brain tissue. Incorporating inferred cortical layer context revealed distinct microglial ligand-receptor associations with ADD in layer 5 involving known AD risk genes, whereas MCI-related patterns were more consistent across layers 1-6. These covariate-adjusted, scRNAseq-based, multiple-sample associations generate layer- and cell-type-specific hypotheses for validation and may ultimately help inform the development of precise therapeutics.

## Data Availability

All data produced in the present study are available upon reasonable request to the authors

https://www.radc.rush.edu

## LIST OF ABBREVIATIONS

ADD: Alzheimer’s disease dementia
snRNAseq: single-nucleus RNA sequencing
CCC: cell-cell communication
MCI: mild cognitive impairment
WM: white matter
DLPFC: dorsolateral prefrontal cortex
ROS: Religious Order Study
MAP: Rush Memory and Aging
LIBD: Lieber Institute for Brain Development
NCI: no cognitive impairment
LDL: low-density lipoprotein
LRPs: low-density lipoprotein receptor-related proteins
SNPs: single-nucleotide polymorphisms
QC: quality control

## ACKNOWLEDGMENTS

This work was supported by National Institutes of Health grant awards R01AG089703 (JY), RF1AG071170 (QD, MPE), and R01MH126449 (MPE). ROSMAP is supported by P30AG10161, P30AG72975, R01AG15819, R01AG17917, U01AG46152, and U01AG61356.

## CONSENT STATEMENT

All ROSMAP participants enrolled without known dementia and agreed to detailed clinical evaluation and brain donation at death. Both studies were approved by an Institutional Review Board of Rush University Medical Center (ROS IRB# L91020181, MAP IRB# L86121802). Both studies were conducted according to the principles expressed in the Declaration of Helsinki. Each participant signed an informed consent, Anatomic Gift Act, and an RADC Repository consent (IRB# L99032481) allowing her data and biospecimens to be repurposed. ROS/MAP transcriptomics data were shared with a data use agreement.

## AUTHOR’S CONTRIBUTION

Q.D. conducted data analysis and drafted the manuscript; M.M. and J.L. conducted data analysis; S.Tang annotated the cortical layer for ROS/MAP data; J.H. supervised the cortical layer annotation prediction; S.Tasaki, Y.W., D.A.B., V.M., P.L.D, and A.S.B. collected the ROS/MAP phenotype and snRNAseq data and participated in data interpretation; M.P.E. and J.Y. conceived this study and supervised data analysis and data interpretation. All authors revised the manuscript.

## DATA AVAILABILITY

ROS/MAP data: www.radc.rush.edu

ROS/MAP snRNAseq data: https://www.synapse.org/Synapse:syn31512863

CelEry tool: https://github.com/QihuangZhang/CeLEry

scater R package: https://bioconductor.org/packages/release/bioc/html/scater.html

scuttle R package: https://www.bioconductor.org/packages/release/bioc/html/scuttle.html

LIANA+ tool: https://github.com/saezlab/liana-py

STACCato tool: https://github.com/daiqile96/STACCato

NHGRI-EBI GWAS Catalog: https://www.ebi.ac.uk/gwas/

Scripts for this study: https://github.com/daiqile96/ROSMAP_C2C

