## SupplementaryInformation for "Cell-Cell Communication Patterns in Alzheimer’s Disease Differ by Cortical Layer"

**Supplementary Figures**

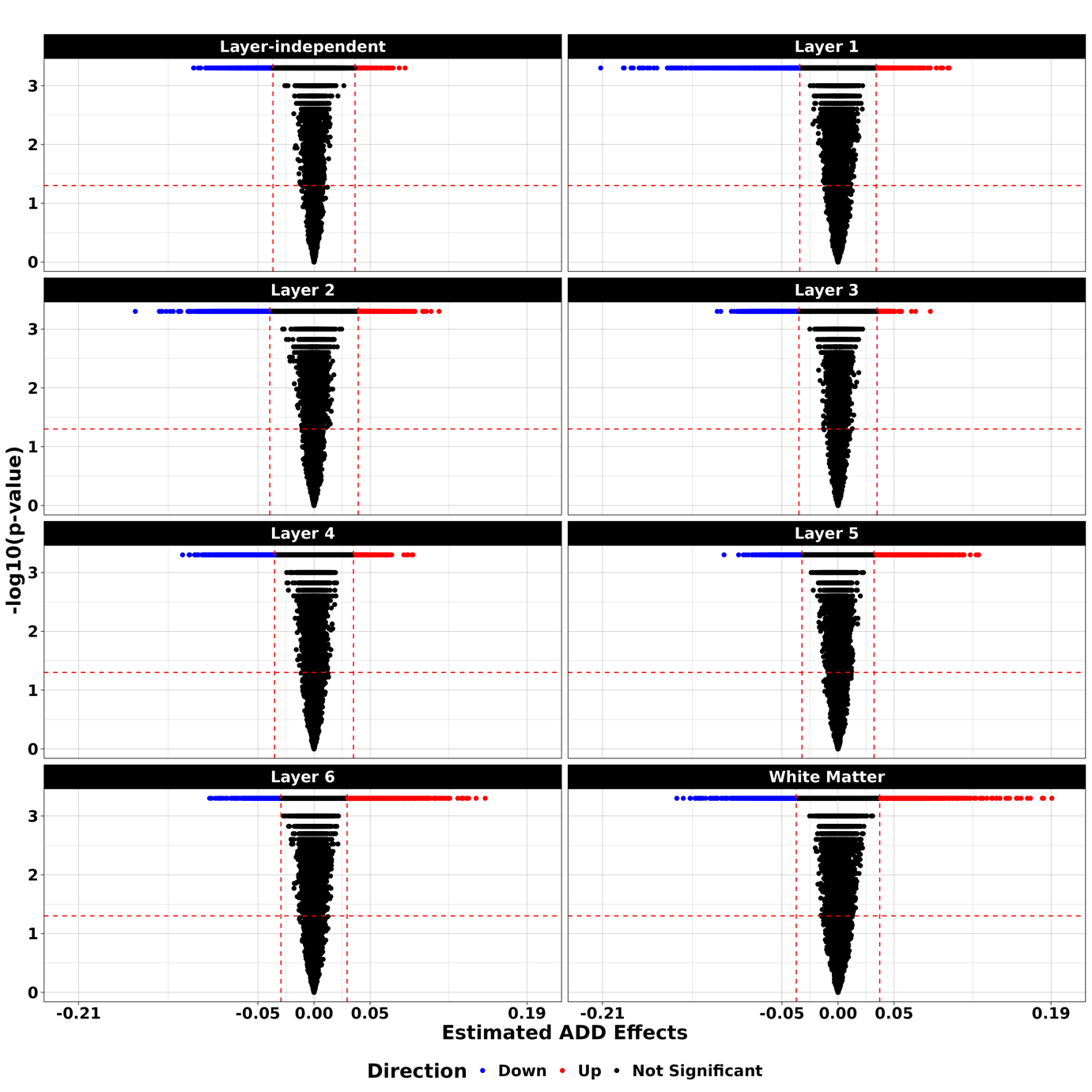
**Figure S1. Significance levels of the estimated ADD effects of all CCC events, generated by 4,999 iterations of bootstrapping resampling.** Putative ADD-related CCC events with p-values < 0.05 and magnitudes of their estimated ADD effects as the top 5% among all CCC events are colored blue for negative effects and red for positive effects. Non-significant ones are shown in black. Y-axis represents -log10(p-values) and x-axis represents the estimated disease effects.

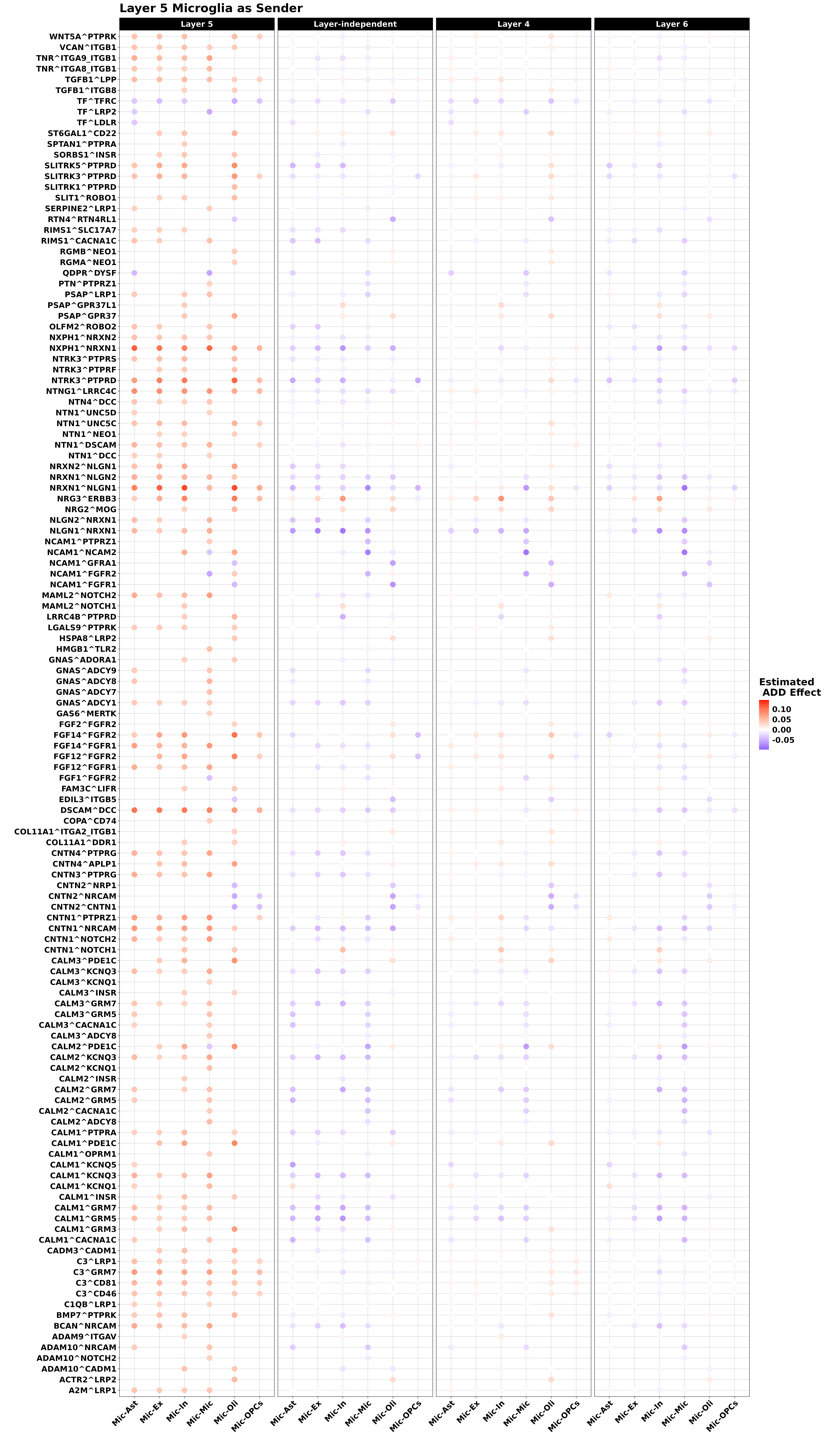
**Figure S2. The estimated ADD disease effects on putative CCC events in Layer 5 results where microglia act as the sender cell type, and the ligand-receptor pairs do not involve genes significantly associated with ADD in the GWAS catalog.** The estimated ADD disease effects for these CCC events are compared between Layer 5 results and layer-independent results, as well as Layer 4 and Layer 6 results. Positive disease effects, shown in red, indicate positive associations between CCC events and ADD, while negative disease effects, shown in blue, indicate negative associations. Within each panel, each grid represents the estimated ADD disease effect for a specific CCC event, with the ligand-receptor pair labeled on the y-axis and the sender-receiver cell type pair indicated on the x-axis. Abbreviations: astrocytes (Ast), excitatory neurons (Ex), inhibitory neurons (In), microglia (Mic), oligodendrocytes (Oli), and oligodendrocyte precursor cells (OPCs), genome-wide association study (GWAS).

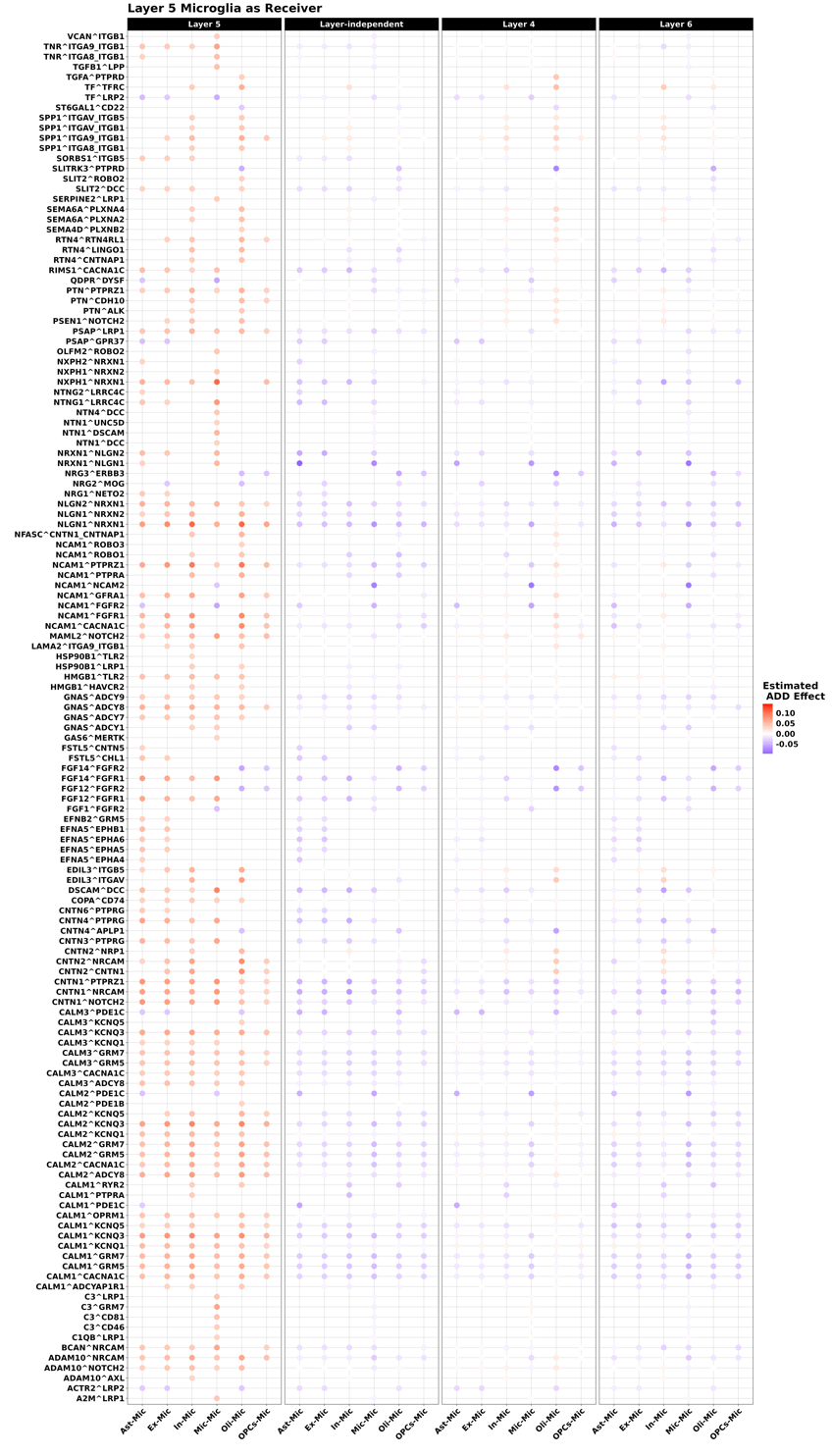
**Figure S3. The estimated ADD disease effects on putative CCC events in Layer 5 results where microglia act as the receiver cell type, and the ligand-receptor pairs do not involve genes significantly associated with ADD in the GWAS catalog.** The estimated ADD disease effects for these CCC events are compared between Layer 5 results and layer-independent results, as well as Layer 4 and Layer 6 results. Positive disease effects, shown in red, indicate positive associations between CCC events and ADD, while negative disease effects, shown in blue, indicate negative associations. Within each panel, each grid represents the estimated ADD disease effect for a specific CCC event, with the ligand-receptor pair labeled on the y-axis and the sender-receiver cell type pair indicated on the x-axis. Abbreviations: astrocytes (Ast), excitatory neurons (Ex), inhibitory neurons (In), microglia (Mic), oligodendrocytes (Oli), and oligodendrocyte precursor cells (OPCs), genome-wide association study (GWAS).

**Figure S4. Significance levels of the estimated MCI effects of all CCC events, generated by 4,999 iterations of bootstrapping resampling.** Detected putative MCI-related CCC events with p-values < 0.05 and magnitudes of their estimated MCI effects as the top 5% among all CCC events are colored blue for negative effects and red for positive effects. Non-significant ones are shown in black. Y-axis represents -log10(p-values) and x-axis represents the estimated disease effects.

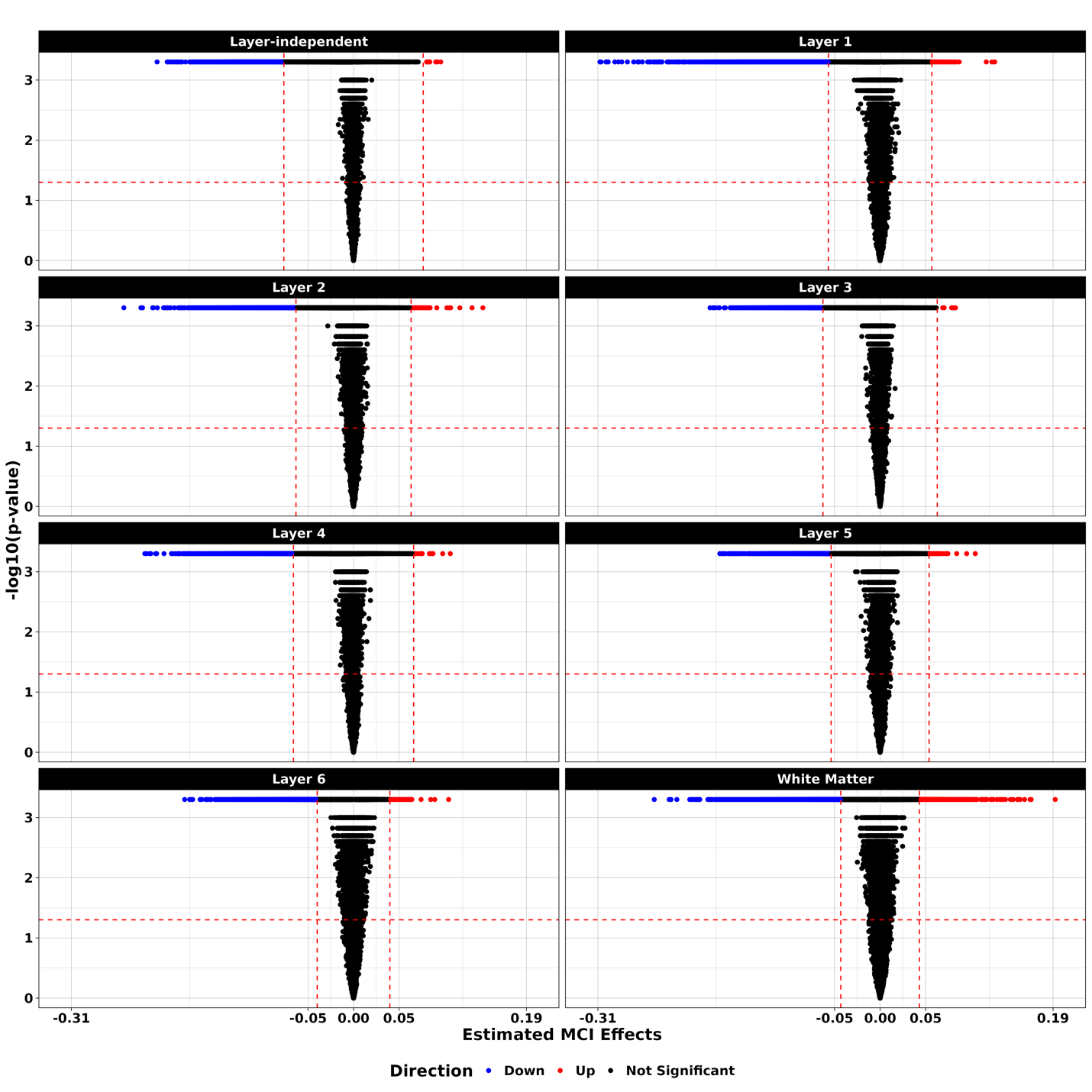

**Figure S5. (A) Bar plot quantifying the Jaccard Index for the overlap of detected putative MCI-related CCC events between layer-independent and each layer-dependent result. (B) Heatmap of the Jaccard Index to show the overlap of MCI-related CCC events between each pair of layer-dependent results.** Abbreviations: white matter (WM).

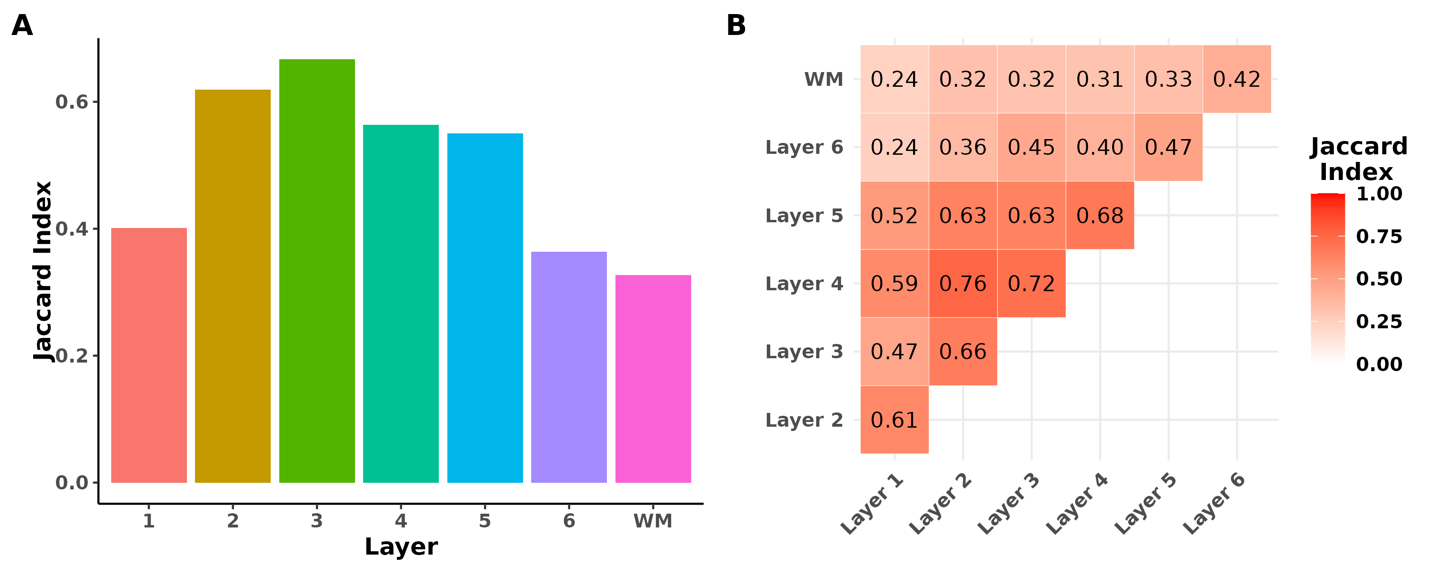

**Figure S6. Circos plots of MCI-related communication network between cell types identified from** (A) layer-independent analysis, (B) Layer 1, (C) Layer 2, (D) Layer 3, (E) Layer 4, (F) Layer 5, (G) Layer 6, and (H) white matter. The width of the links represents the total number of putative MCI -related CCC events between each pair of sender-receiver cell types. The number of identified positive MCI-related CCC events is shown on the left side of each panel, while the negative events are on the right. Each color represents a different cell type.

Abbreviations: astrocytes (Ast), excitatory neurons (Ex), inhibitory neurons (In), microglia (Mic), oligodendrocytes (Oli), and oligodendrocyte precursor cells (OPCs).

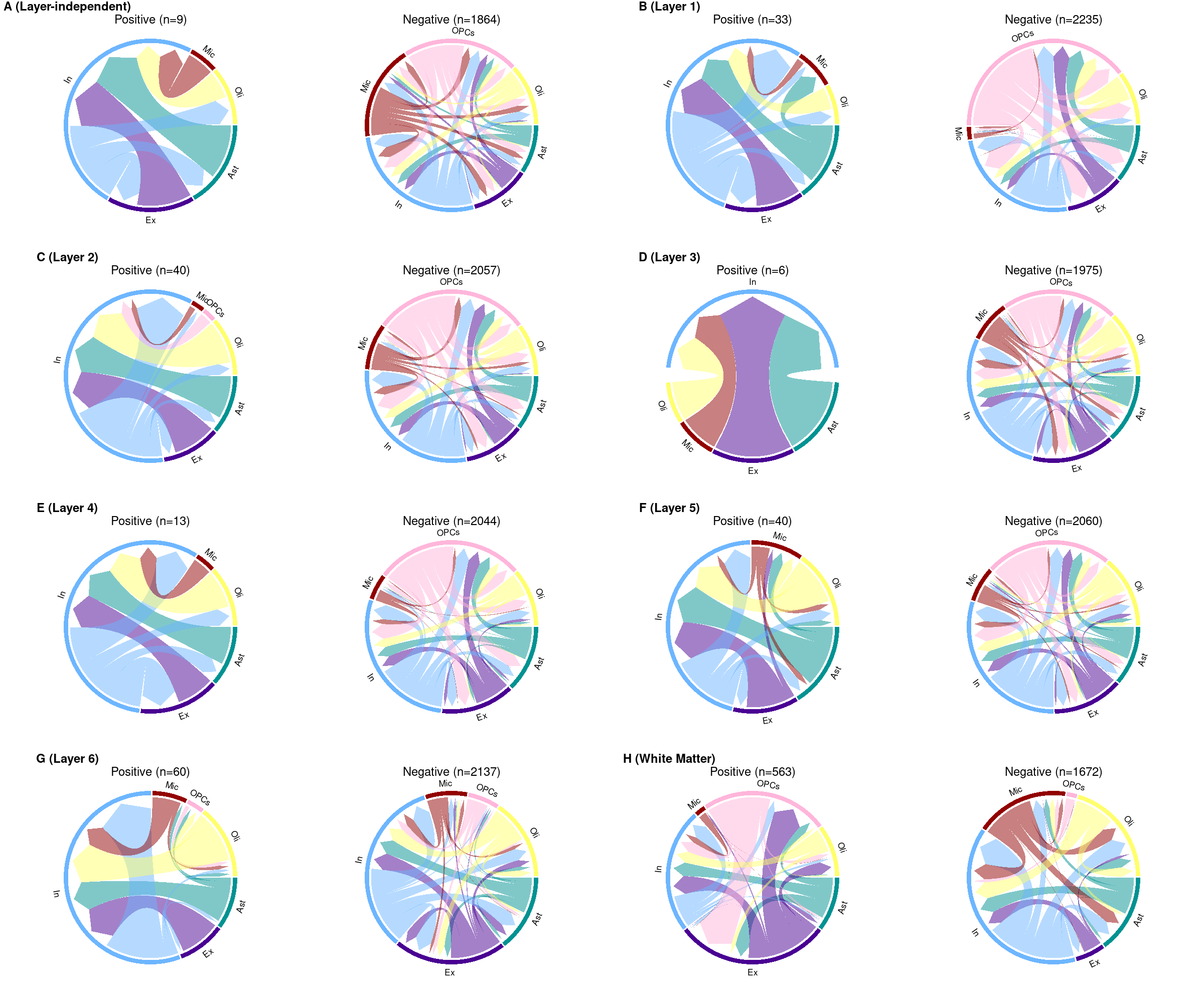

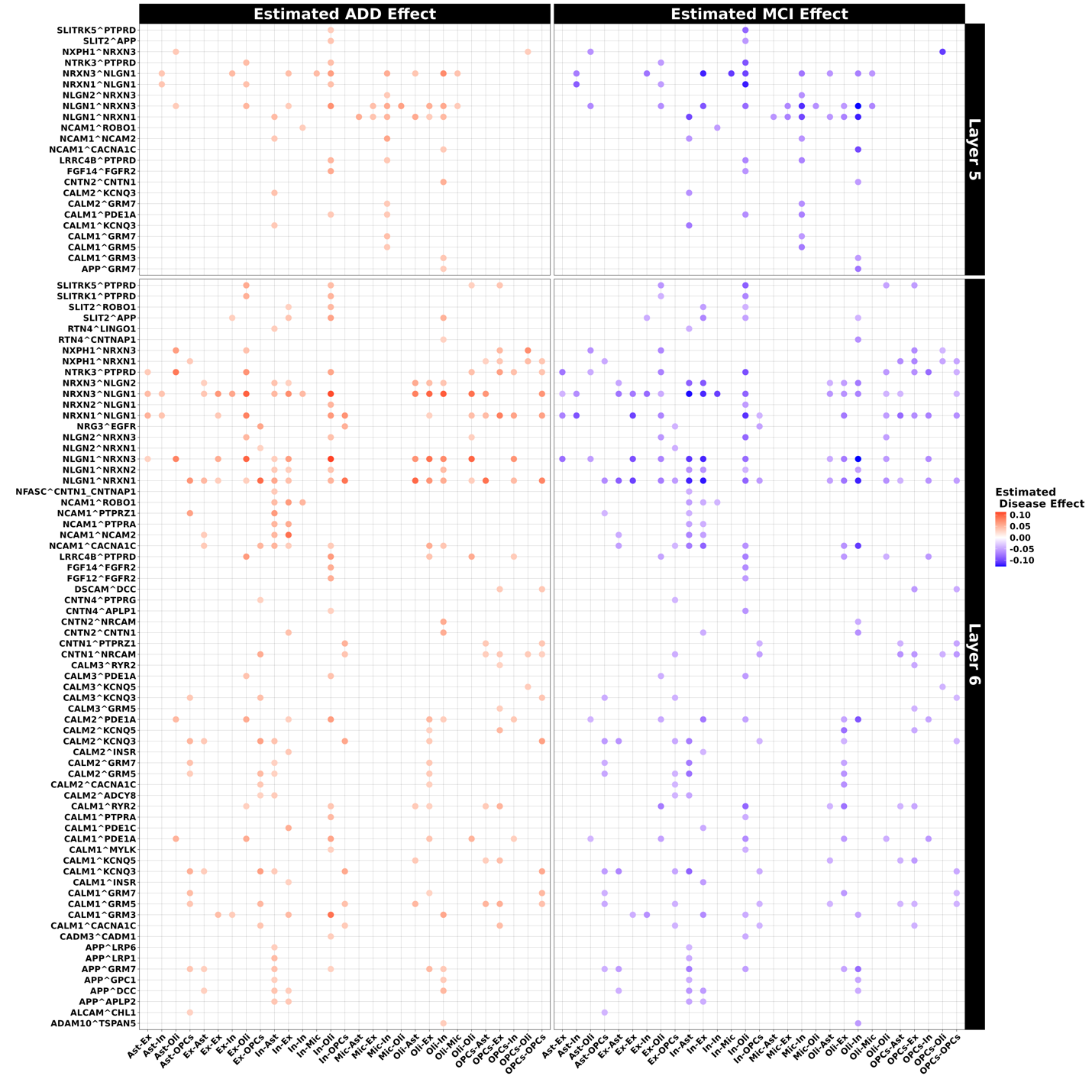
**Figure S7. The estimated ADD and MCI disease effects on putative CCC events with opposite directions between ADD and MCI effects in Layer 5 and Layer 6.** The left panel shows the ADD effects, and the right panel shows the MCI effects. The top panel shows the Layer 5 results, and the bottom panel shows the Layer 6 results. Positive disease effects, shown in red, indicate positive associations between CCC events and ADD, while negative disease effects, shown in blue, indicate negative associations. Within each panel, each grid represents the estimated effect for a specific CCC event, with the ligand-receptor pair labeled on the y-axis and the sender-receiver cell type pair indicated on the x-axis.

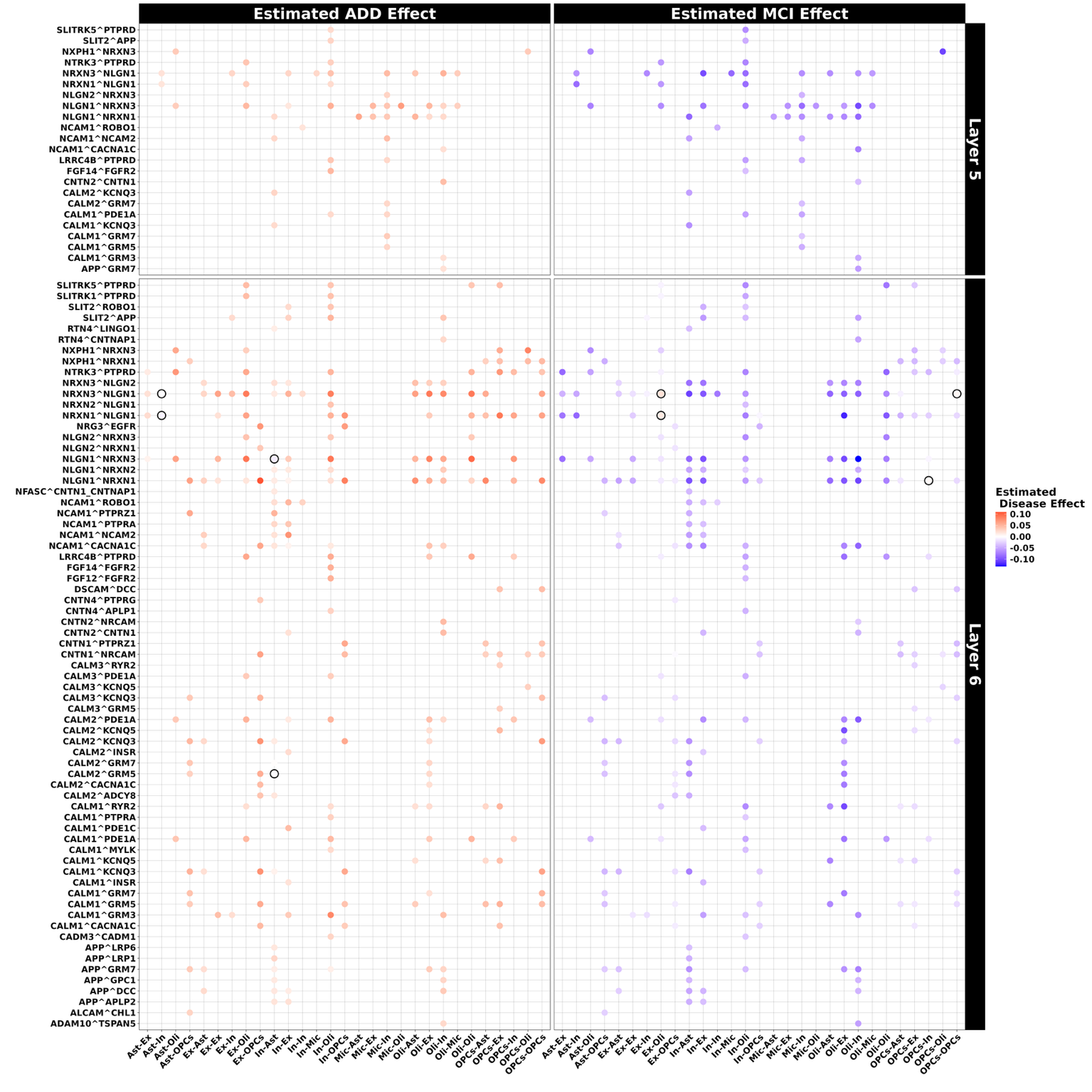
**Figure S8. The estimated ADD and MCI disease effects by the sub-cohort analyses for the putative ADD/MCI related CCC events with opposite ADD and MCI effect sizes detected by the full-cohort analyses in Layer 5 and Layer 6.** The left panel shows the ADD effects, and the right panel shows the MCI effects. The top panel shows the Layer 5 results, and the bottom panel shows the Layer 6 results. Positive disease effects, shown in red, indicate positive associations between CCC events and ADD, while negative disease effects, shown in blue, indicate negative associations. Within each panel, each grid represents the estimated effect for a specific CCC event, with the ligand-receptor pair labeled on the y-axis and the sender-receiver cell type pair indicated on the x-axis. The circles represent the CCC events with flipped effect size directions by sub-cohort analyses.

**Supplementary Table**

|  | **# of Samples** | **# of ADD** | **# of MCI** | **# of NCI** | **# of Sender & Receiver Cell Types** | **# of Ligand-Receptor Pairs** | **# of Putative ADD/MCI Related CCC events (Top 5%)** |
| --- | --- | --- | --- | --- | --- | --- | --- |
| **Whole ROS/MAP** | 395 | 150 | 107 | 138 | 6 | 1,041 | 1,873 |
| **Layer 1** | 388 | 146 | 105 | 137 | 6 | 1,260 | 2,268 |
| **Layer 2** | 395 | 150 | 107 | 138 | 6 | 1,165 | 2,097 |
| **Layer 3** | 395 | 150 | 107 | 138 | 6 | 1,101 | 1,981 |
| **Layer 4** | 395 | 150 | 107 | 138 | 6 | 1,143 | 2,057 |
| **Layer 5** | 390 | 147 | 105 | 138 | 6 | 1,167 | 2,100 |
| **Layer 6** | 388 | 146 | 105 | 137 | 6 | 1,221 | 2,197 |
| **WM** | 383 | 145 | 104 | 134 | 6 | 1,242 | 2,235 |

**Table S1.** The numbers of samples of ADD, MCI, NCI statuses, considered cell type numbers, ligand-receptor pair numbers, and the total number of putative ADD/MCI related CCC events detected by STACCato were presented.

**Supplementary Methods**

### **STACCato analysis of ADD- and MCI- related CCC events**

In this study, we aimed at identifying CCC events associated with ADD or MCI within each cortical layer (i.e., by layer-dependent analyses), and then compared the findings to those obtained without accounting for spatial context (i.e., layer-independent analysis). Layer-dependent analyses were conducted by analyzing nuclei that were annotated each target cortical layer (or white matter), whereas the layer-independent analyses were conducted by using all nuclei in all six cortical layers and white matter. The same data preprocessing and CCC analysis procedures were applied to the subsets of snRNAseq data of Layers 1-6 and WM, as well as the whole ROS/MAP snRNAseq dataset. We considered 6 major brain cell types –– astrocytes, excitatory neurons, inhibitory neurons, microglia, oligodendrocytes, and oligodendrocyte precursor cells. For each of these CCC analyses, we employed our recently developed tool, STACCato^1^, implementing a tensor regression model to identify ADD-related and MCI-related CCC events, while adjusting for potential confounders, including age, gender, education, *APOE* E4 and *TOMM40* genetic variants, and recent medication usages.

#### **snRNAseq data QC and preprocessing**

For each of the corresponding ROS/MAP RNAseq dataset (per cortical layer, or white matter, or the whole prefrontal cortex), we first conducted quality control (QC) by filtering out low-quality cells. Cells expressing fewer than 200 genes were excluded. Mitochondrial genes were identified by their names, and cells with mitochondrial gene expression exceeding 15% were removed to eliminate potentially unhealthy cells. Additionally, genes detected in fewer than 3 cells were filtered out. Next, we filtered samples based on the number of cell types. Samples with fewer than 2 unique cell types were excluded, as the subsequent CCC analysis focuses on communications between cell types.

Table S1 summarizes the number of samples retained after data quality control for each dataset. For the layer-independent analysis, all 395 samples with complete data of the summary clinical cognitive diagnoses and considered covariates. Analyses of Layers 2, 3, and 4 also included all 395 samples. Analyses of Layers 1, 5, 6, and white matter excluded 7, 5, 7, and 12 samples, respectively, as these samples contained only a single cell type. The numbers of ADD, MCI, and NCI samples included in the analyses for these layers are detailed in Table S1.

The snRNAseq data after QC were quantified as pseudo bulk read counts per gene per sample, normalized by scaling library size factors to ${10}^{4}$, and then subjected to log-transformation to calculate CCC scores for follow-up analyses by STACCato. These QC and preprocessing steps were performed using the *SingleCellExperiment*^2^, *scater*^3^, and *scuttle*^3^ packages in R software.

#### **Construct CCC scores using LIANA+**

For each of the CCC analyses, we first used LIANA+^4^ in R software to perform CCC inference based on log-transformed pseudo-bulk expression data. LIANA+ utilizes snRNAseq expression data to quantify the communication strengths of ligand-receptor pairs between each combination of sender and receiver cell type. Specifically, for the CCC event in sample $i$ involving the interaction of ligand-receptor pair $j$ from sender cell type $k$ to receiver cell type $l,$ LIANA+ generates the following communication score $y_{ijkl}$:

$y_{ijkl}= f\left( \mathrm{ligand}_{ijk}, \mathrm{receptor}_{ijl} \right).$ (***Equation S1***)

Here, $\mathrm{ligand}_{ijk}$ denotes the expression of the ligand gene of the ligand-receptor pair $j$ in sender cell type $k$ of sample $i$; $\mathrm{receptor}_{ijl}$ denotes the expression of the receptor gene of the ligand-receptor pair $j$ in receiver cell type $l$ of sample $i$; and $f$ denotes the scoring function. LIANA+ provides multiple choices of the scoring functions. We utilized the mean scoring function $y_{ijkl}=\frac{\mathrm{ligand}_{ijk}+ \mathrm{receptor}_{ijl}}{2}$proposed by CellPhoneDB^5^, along with the consensus list of ligand-receptor pairs provided in the LIANA+ package. These calculated communication scores represent the inferred strengths of CCC events. For each sample in the snRNAseq data, we calculated communication scores for all CCC events across all ligand-receptor pairs from the consensus list, as well as all sender and receiver cell type combinations. These scores from all samples were then arranged into a 4-dimensional tensor with dimensions representing samples, ligand-receptor pairs, sender cell types, and receiver cell types.

We also filtered ligand-receptor pairs at both the sample and dataset levels when applying LIANA+. For each sample, only ligand-receptor pairs expressed in more than 10% of cells were retained to calculate communication scores. When constructing the communication score tensor across all samples, we applied an additional criterion where we retained only ligand-receptor pairs present in more than one-third of the samples. This filtering resulted in approximately 1,000 to 1,200 ligand-receptor pairs per dataset (Table S1). We noted that datasets with larger numbers of cells and samples underwent stricter selection criteria, which reduced the number of retained ligand-receptor pairs. For instance, the layer-independent analysis with 395 samples retained only 1,041 ligand-receptor pairs, while the sub-dataset of white matter with 383 samples retained 1,242 ligand-receptor pairs.

#### **Detect ADD/MCI-related CCC events by STACCato.**

Using the cell-cell communication scores generated by LIANA+, we applied STACCato to detect CCC events associated with ADD or MCI. For each CCC event, STACCato estimates the effect of cognitive status on the communication scores and provides the corresponding p-value to determine if the studying CCC event is significantly influenced by cognitive status. Specifically, STACCato considers the following regression model:

$$y_{ijkl}=\beta_{1,jkl}x_{i,1}+\beta_{2,jkl}x_{i,2}\cdots+\beta_{Q,jkl}x_{i,Q} + \epsilon_{ijkl};$$

$i=1, \cdots I; j=1, \cdots J; k=1,\cdots K; l=1,\cdots L;$(***Equation S2***)

where $x_{i,1}, x_{i,2}, \ldots, x_{i,Q}$ denote *Q* sample-level variables collected for each sample $i$, including the cognitive status and additional covaries.

The above regression model (***Equation S2***) aims to estimate the coefficients {$\beta_{q,jkl}, q=1,\ldots, Q\}$, which denote the effects of sample-level variables on the communication scores of the CCC event ($y_{.jkl}$). For example, we can set $x_{i,1}$ = 0 and $x_{i,2}=0$ for NCI, $x_{i,1}$ = 1 and $x_{i,2}=0$ for MCI, and $x_{i,1}$ = 0 and $x_{i,2}=1$ for ADD, according to the summary clinical cognitive status of sample $i$. Then CCC events with significant $\beta_{1,jkl}$ are the CCC events affected by MCI, and significant $\beta_{2,jkl}$ are the CCC events affected by ADD. The larger the magnitude of the coefficients, the greater the difference in communication magnitudes between the ADD/MCI and NCI subjects. We refer to CCC events with $\beta_{*,jkl}$ significantly different from 0 as CCC events related to the corresponding cognitive status, while adjusting for additional confounding covariates.

Instead of fitting the regression model (***Equation S2***) per CCC event, to effectively account for interdependencies among CCC events, STACCato employs a tensor-based regression model as follows to enable simultaneous analyze analyses of all CCC events:

$\mathcal{Y=B}\times_{1}\boldsymbol{X+ℇ;}$	(***Equation S3***)

where $\mathcal{Y}\boldsymbol{\in}\mathbb{R}^{I\times J\times K\times L}$denotes the 4-dimensional communication score tensor with the $(i, j, k, l )$ entry corresponding to $y_{ijkl}$ in ***Equation S2***;$\boldsymbol{X\in}\mathbb{R}^{I \times Q}$ denotes sample-level design matrix of $Q$ variables; $\times_{1}$ denotes multiplying a tensor by a matrix in the tensor’s first dimension; $\boldsymbol{ℇ\in}\mathbb{R}^{I\times J\times K\times L}$ denotes a 4-dimensional tensor with the $(i, j, k, l )$ entry corresponding to $\epsilon_{ijkl}$ in ***Equation S2***.

We applied STACCato to fit a tensor-based regression model (***Equation S3***) with the constructed 4-dimensional communication score tensor as the outcome variable, and sample-level variables including ADD status, MCI status, age at death, gender, education levels, *TOMM40* haplotypes, *APOE* genotypes, and recent medication usages (including ADD medications, lipid-lowering medications, and urinary condition medications) as independent variables. This framework allowed us to estimate the effects of ADD and MCI on CCC events while accounting for additional covariates. We selected those CCC events with p-values < 0.05 and effect size magnitudes ranking within the top 5% for ADD or MCI, respectively, as the ADD/MCI related CCC events.
